# Determinants of Covid-19 vaccine uptake among the elderly aged 58 years and above in Kericho County, Kenya: Institution based cross sectional survey

**DOI:** 10.1101/2023.01.16.23284598

**Authors:** Calvince Otieno Anino, Immaculate Wandera, Zachary Masimba Ondicho, Collins Kipruto Kirui, Carjetine Syallow Makero, Phanice Kerubo Omari, Philip Sanga

## Abstract

**Background:** Hesitancy to Covid-19 vaccine is a global challenge despite the compelling evidence of the value of vaccine in preventing disease and saving lives. It is suggested that context-specific strategies can enhance acceptability and decrease hesitancy to Covid-19 vaccine. Hence, the study determined uptake and determinants of Covid-19 vaccine following a sustained voluntary vaccination drive by Kenyan government.

**Method:** We conducted institution based cross-sectional survey of 1244 elderly persons aged 58 to 98 years in the months of January, February and March, 2022. A multinomial logistic regression analysis was used to investigate determinants of Covid 19 vaccine uptake. The predictor variables included socioeconomic and demographic characteristics, convenience and ease of access of the vaccine, collective responsibility, complacency and the three dimensions of confidence; trust in safety, trust in decision makers and delivery system. The findings are reported as the adjusted odd ratio (AOR) at 95% confidence interval (CI). Significant level was considered at p <0.05.

**Result:** The results from the multinomial logistic regression analysis indicated that advanced age and presence of chronic disease were associated with increased odds of doubt on Covid 19 vaccine, while long distance from vaccination centers was associated with increased odds of delay in vaccination.

**Conclusion:** Overall, the findings of this study have provided valuable insights into the factors influencing vaccine hesitancy among the elderly population in Kenya and will inform the development of targeted interventions to increase vaccine acceptance and uptake in this population.

## Background

Covid-19 vaccine hesitancy has become a significant global challenge in the effort to control the spread of the pandemic (1). Despite the overwhelming evidence of the efficacy and safety of vaccines in preventing disease and saving lives (2), some individuals and communities have expressed skepticism or refusal to receive the vaccine. This hesitancy can have serious consequences, as it can lead to decreased vaccine uptake and ultimately contribute to the continued spread of the disease (3, 4). To address this issue, it is important to understand the prevalence and determinants of Covid-19 vaccine hesitancy (5). A range of factors have been identified as contributing to vaccine hesitancy, including socioeconomic and demographic characteristics, awareness and knowledge of the vaccine, attitudes towards collective responsibility, complacency, and confidence in the vaccine and vaccination process (6, 7). Targeted interventions, such as education campaigns and addressing misinformation, have been shown to be effective in reducing vaccine hesitancy in some populations (8, 9). However, it is essential to recognize that these interventions may not be equally effective in all contexts and that it is necessary to examine the specific factors driving hesitancy in different populations (10, 11). In Kenya, the government has implemented a sustained voluntary vaccination drive as part of its efforts to control the spread of Covid-19. Despite this, little is known about the prevalence and determinants of vaccine hesitancy in this population. The current study aims to fill this gap by conducting a cross-sectional survey of elderly individuals aged 58 to 98 years in Kenya to investigate the prevalence and determinants of Covid-19 vaccine hesitancy. The results of this study will provide valuable insights into the specific factors driving vaccine hesitancy in this population and inform the development of context-specific strategies to increase vaccine acceptability and uptake.

## Methods

### Study design, sampling procedure and ethical consideration

In order to investigate the prevalence and determinants of Covid-19 vaccine hesitancy in Kenya, we conducted an institution-based cross-sectional survey of 1244 elderly individuals aged 58 to 98 years in three sub-counties of Kericho County in the Southern Rift Valley of Kenya. These sub-counties were purposively chosen as they have the highest proportion of elderly individuals in the county and together account for over 50% of the county’s population. Stratified sampling was used to select 20 health facilities, including 12 government and 8 private facilities, that offer Covid-19 vaccination. Systematic random sampling was then used to select 63 respondents from each of these facilities. The questionnaires were administered in private rooms by trained interviewers, who were graduate public health students on their 10th month of internship and had knowledge of the 5Cs of vaccine hesitancy (confidence, complacency, convenience, and collective responsibility). The interviews were conducted in a confidential manner, with coded lists used to assign unique codes to each respondent in order to maintain their anonymity. Tablet computers with the open data kit collect application were used by the interviewers to collect the data, which was then submitted to a central server at the end of each day. This study was approved by the University of Kabianga Institutional Research Ethics Committee and written informed consent was obtained from each respondent.

### Study variables

#### Demographic characteristics

In order to gather demographic information, we collected data on participants’ age, gender, marital status, level of education, and occupation. Age was recorded as a continuous variable but was later transformed into a categorical variable for analysis, with the categories being under 70 years, 70 to 80 years, and above 80 years. We also asked participants about their primary occupation and whether they had any history of chronic disease or had taken any vaccine in the last five years. In addition, we recorded whether participants were healthcare workers and their area of residence. All of this information was collected using yes or no responses.

#### Covid 19 vaccine uptake categories

The primary focus of this study was to assess the determinants of Covid-19 vaccine uptake. To do this, we first asked participants whether they were aware of the Covid-19 vaccine. We then measured vaccine uptake by asking participants if they had received any of the Covid-19 vaccines. We used a single item measure with four possible responses: (1) accepting the vaccine without doubt for reasons other than allergies or illness, (2) accepting the vaccine with doubt for reasons other than allergies or illness, (3) refusing the vaccine for reasons other than allergies or illness, or delaying the vaccine for reasons other than allergies or illness. Based on their responses, participants were classified into one of four categories: refusers, delayers, acceptors with doubt, or no vaccine hesitancy. We used the classification criteria defined by (12) in order to classify vaccine acceptance and hesitancy.

### Determinants of Covid 19 vaccine uptake

To assess the determinants of Covid-19 vaccine uptake, we used a modified version of the 5C model of the psychological antecedents to vaccination (2). This model suggests that complacency, constraints, confidence, collective responsibility, and calculation are important predictors of vaccine hesitancy. We assessed confidence in three dimensions: trust in the safety and effectiveness of the Covid-19 vaccines, trust in the government officials who make decisions about the Covid-19 vaccines, and trust in the delivery of the Covid-19 vaccination services with regards to the competency and reliability of the healthcare workers. We used a 10-item scale with a 5-point hedonic response scale to measure the extent to which participants agreed with these dimensions of confidence. The variables measured in the dimension of trust in vaccine safety included concerns about safety, unknown side effects, long-term effects, harmful substances in the vaccine, and the short development time of the vaccine. We also assessed trust in the safety and effectiveness of the vaccine with regard to religion compatibility and used a scoring system with responses ranging from 1 (strongly disagree) to 5 (strongly agree). Trust in the delivery of the Covid-19 vaccination services was measured using a similar scoring system, with responses ranging from 1 (strong distrust) to 5 (strong trust) for vaccine manufacturers, professional institutions, and healthcare providers. We also used a similar scoring system to assess trust in decision-makers, including government officials, politicians, and church leaders. For the purpose of analysis, responses to each of the 10 items were further classified into two categories: agree (including responses of “strongly agree” and “agree” for trust in vaccines, and “strong trust” and “trust” for the Covid-19 vaccination delivery system and decision-makers) and disagree (including responses of “strongly disagree” and “disagree” for trust in vaccines, and “strong distrust” and “distrust” for the Covid-19 vaccination delivery system and decision-makers).

We also measured collective responsibility using a 3-point scale, with responses of “always,” “sometimes,” and “never” to questions about mask-wearing, physical distancing, and hand hygiene in public and at home. In addition, we assessed the importance, information, opinion, advice, and beliefs of participants as determinants of Covid-19 vaccine uptake. Some of the questions asked included: “Do you know much information about the Covid-19 vaccine?” “Is the opinion of your family and friends important to your decision to take or not take the Covid-19 vaccine?” “Do you value the advice of health professionals regarding the effectiveness of the Covid-19 vaccine?” and “Do you believe that the Covid-19 vaccine will help protect the people who take it?” These variables were used to understand the various factors that may influence an individual’s decision to receive or not receive the Covid-19 vaccine.

### Data analysis

To analyse the data collected from this study, we used multinomial logistic regression analysis to investigate the prevalence and determinants of Covid-19 vaccine hesitancy in the elderly population in Kenya. The predictor variables, or potential determinants of vaccine hesitancy, were socioeconomic and demographic characteristics, awareness of the Covid-19 vaccine, attitudes towards collective responsibility, complacency, and the three dimensions of confidence. The dependent variable was the vaccine hesitancy status of each participant, as classified into one of four categories: refusers, delayers, acceptors with doubt, or no vaccine hesitancy.

We have presented the findings as adjusted odds ratios (AOR) at a 95% confidence interval (CI). A p-value of less than 0.05 was considered statistically significant. We also conducted subgroup analyses to examine any potential differences in vaccine hesitancy among different demographic subgroups. Additionally, we used descriptive statistics to summarize the sociodemographic characteristics of the study sample and to provide an overview of the prevalence of vaccine hesitancy among the elderly population in Kenya. We also used cross-tabulations and chi-square tests to explore any potential associations between vaccine hesitancy and the various predictor variables included in the analysis.

## Results

### Socio-demographic characteristics and vaccine uptake

The results of this study show that, among the elderly population in Kenya, a significant proportion expressed hesitancy towards the Covid-19 vaccine. Of the respondents, 81.5% were aware of the vaccine, but only 27.4% accepted it without any doubts, while 14.5% accepted it with doubt, 37.1% were delayers, and 21% were refusers.

In terms of sociodemographic characteristics, all variables, except for gender and county of residence, were significantly associated with vaccine uptake. Those who were married or aged below 70 years were more likely to be classified as acceptors with doubts, delayers, or have an intention to refuse the vaccine, compared to their respective cohort categories. Additionally, those with secondary or post-secondary education were more likely to be classified as acceptors with doubts, and a significant proportion of farmers had an intention to refuse the vaccine.

### Association of confidence dimensions, collective responsibility, convenience and complacency with Covid 19 uptake

Table 2 and 3 show the association of confidence dimensions, collective responsibility, and complacency with Covid-19 vaccine uptake. All three dimensions of confidence were significantly associated with vaccine uptake. Trust in vaccine safety was significantly associated with no hesitancy (87.7%), while trust in the delivery system was significantly associated with no hesitancy (94.7%) and acceptance with doubts (63.9%). Specific trust parameters, such as trust in the ability of the vaccine to protect, were also significantly associated with no hesitancy and acceptance with doubts. However, concerns about long-term effects and unknown side effects were highly associated with acceptance with doubts, delay, and refusal.

**Table 1.** Socio-demographic characteristics of the respondents and Covid 19 vaccine uptake level

| Variables | n (%) | Covid 19 vaccine uptake |  |  |  | P-value |
| --- | --- | --- | --- | --- | --- | --- |
|  |  | No hesitancy | Acceptors with doubts | Delayers | Refusers |  |
|  | **1244** | **341 (27.4)** | **180 (14.5%)** | **462 (37.1%)** | **261 (21%)** |  |
| Gender |  |  |  |  |  |  |
| Male | 692 (55.6%) | 164 (48.1%) | 106 (58.9%) | 297 (64.3%) | 125 (47.9%) | 0.63 |
| Female | 552 (44.4%) | 177 (51.9%) | 74 (41.1%) | 165 (35.7%) | 136 (52.1%) |  |
| Age category |  |  |  |  |  |  |
| <70 years | 913 (73.4%) | 236 (69.2%) | 139 (77.2%) | 354 (76.6%) | 184 (70.5%) | 0.01* |
| 70 to 80 years | 281 (22.6%) | 91 (26.7%) | 34 (18.9%) | 97 (21%) | 59 (22.6%) |  |
| >80 years | 50 (4%) | 14 (4.1%) | 7 (3.9%) | 11 (2.4%) | 18 (6.9%) |  |
| Marital status |  |  |  |  |  |  |
| Single | 30 (2.4%) | 6 (1.8%) | 13 (7.2%) | 6 (1.3%) | 5 (1.9%) | <0.01** |
| Married | 1194 (96%) | 333 (97.6%) | 159 (88.3%) | 448 (97%) | 254 (97.3%) |  |
| Divorced | 20 (1.6%) | 2 (0.6%) | 8 (4.5%) | 8 (1.7%) | 2 (0.8%) |  |
| Level of education |  |  |  |  |  |  |
| Primary | 478 (38.4%) | 158 (46.3%) | 13 (7.2%) | 196 (42.4%) | 111 (42.5%) | 0.02* |
| Secondary | 295 (23.7%) | 44 (12.9%) | 116 (64.4%) | 88 (19%) | 47 (18%) |  |
| Post-secondary | 214 (17.2%) | 123 (36.1%) | 43 (23.9%) | 20 (4.3%) | 28 (10.7%) |  |
| None | 257 (20.7%) | 16 (4.7%) | 8 (4.4%) | 158 (34.2%) | 75 (28.7%) |  |
| Primary occupation |  |  |  |  |  |  |
| Farming | 766 (61.6%) | 139 (40.8%) | 94 (52.2%) | 298 (64.5%) | 235 (90%) | 0.03* |
| Business | 175 (14.1%) | 43 (12.6%) | 61 (38.9%) | 65 (14.1%) | 6 (2.3%) |  |
| Informal | 303 (24.4%) | 159 (46.6%) | 25 (13.9%) | 99 (21.4%) | 20 (7.7%) |  |
| Residence |  |  |  |  |  |  |
| Kericho County | 953 (76.6%) | 259 (76%) | 121 (67.2%) | 389 (84.2%) | 184 (70.5%) | 0.88 |
| Other counties | 291 (23.4%) | 82 (24%) | 59 (32.8%) | 73 (15.8%) | 77 (29.5%) |  |
| Healthcare worker |  |  |  |  |  |  |
| Yes | 58 (4.7%) | 43 (12.6%) | 9 (5%) | 4 (0.9%) | 2 (0.8%) | 0.61 |
| No | 1186 (95.3%) | 298 (87.4%) | 171 (95%) | 458 (99.1%) | 259 (99.2%) |  |
| Chronic disease |  |  |  |  |  |  |
| Yes | 682 (54.8%) | 218 (63.9%) | 24 (13.3%) | 302 (65.4%) | 138 (52.9%) | <0.01** |
| No | 562 (45.2%) | 123 (36.1%) | 156 (86.7%) | 160 (34.6%) | 123 (47.1%) |  |
| Awareness |  |  |  |  |  |  |
| Yes | 1014 (81.5%) | 341 (100%) | 180 (100%) | 317 (68.6%) | 176 (67.4%) |  |
| No | 230 (18.5%) |  |  | 145 (31.4%) | 85 (32.6%) |  |

\* Indicates statistical significance ( $p < 0.05$ )

**Table 2:** Association between confidence dimension and Covid 19 uptake

| Variables | Covid 19 vaccine uptake |  |  |  | P-value |
| --- | --- | --- | --- | --- | --- |
|  | No hesitancy<br>341 (27.4%) | Acceptors with doubts<br>180 (14.5%) | Delayers<br>462 (37.1%) | Refusers<br>261 (21%) |  |
| Safety |  |  |  |  |  |
| Vaccine is effective | 93 (27.3%) | 146 (81.1%) | 238 (51.5%) | 71 (27.2%) | 0.03* |
| Unknown side effect | 124 (36.4%) | 150 (83.3%) | 420 (90.9%) | 251 (96.2%) | 0.01* |
| Long term effects | 143 (41.9%) | 134 (74.4%) | 401 (86.8%) | 247 (94.6%) | 0.01* |
| Harmful substance | 57 (16.7%) | 37 (20.6%) | 86 (18.6%) | 54 (20.7%) | 0.25 |
| Too short time for development and testing | 32 (9.4%) | 66 (36.7%) | 194 (42%) | 196 (75.1%) | 0.02* |
| Vaccine is compatible with personal beliefs | 96 (28.2%) | 112 (62.2%) | 237 (51.3%) | 74 (28.4%) | 3.41 |
| Vaccine is compatible with natural remedies | 85 (25%) | 107 (59.4%) | 221 (47.8%) | 65 (24.9%) | 0.72 |
| Covid 19 vaccine completely protect people who take it | 270 (79.2%) | 126 (70%) | 193 (41.7%) | 60 (23%) | 0.01* |
| I/someone I know had a bad experience with previous vaccine | 197 (57.7%) | 12 (6.7%) | 22 (4.8%) | 24 (9.2%) | 0.07 |
| I/someone I know had bad experience with Covid 19 vaccine | 13 (4%) | 104 (57.8%) | 237 (51.3%) | 151 (57.9%) | 0.02* |
| Trust vaccine is safe for use |  |  |  |  |  |
| Agree | 299 (87.7%) | 58 (32.2%) | 124 (26.8%) | 20 (7.7%) | <0.01** |
| Disagree | 42 (12.3%) | 122 (67.8%) | 338 (73.2%) | 241 (92.3%) |  |
| Decision makers |  |  |  |  |  |
| Government officers | 328 (96.2%) | 151 (83.9%) | 375 (81.2%) | 112 (42.9%) | 0.01* |
| Church leaders | 337 (98.8%) | 163 (90.5%) | 413 (89.4%) | 194 (74.3%) | 0.01* |
| Politicians | 114 (33.4%) | 79 (43.9%) | 262 (56.7%) | 223 (85.4%) | <0.01** |
| Trust in decision makers |  |  |  |  |  |
| Agree | 329 (96.5%) | 170 (94.5%) | 431 (93.3%) | 341 (92.3%) | 0.04* |
| Disagree | 12 (3.5%) | 11 (6.5%) | 32 (6.7%) | 20 (7.7%) |  |
| Delivery system |  |  |  |  |  |
| Healthcare workers | 309 (90.6%) | 160 (88.9%) | 388 (84%) | 123 (47.1%) | 0.01* |
| Hospital and other professional institutions | 314 (92.1%) | 167 (92.8%) | 433 (93.7%) | 189 (72.4%) | 0.03* |
| Vaccine manufacture and companies | 292 (85.6%) | 126 (70%) | 289 (62.6%) | 135 (51.7%) | 0.01* |
| Trust in delivery system |  |  |  |  |  |
| Agree | 323 (94.7%) | 115 (63.9%) | 207 (44.8%) | 90 (34.5%) | 0.01* |
| Disagree | 18 (5.3%) | 65 (36.1%) | 255 (55.2%) | 171 (65.5%) |  |

\* Indicates statistical significance ( $p < 0.05$ )

**Table 3.** Association of collective responsibility, convenience and complacency with Covid 19 vaccine uptake

| Variables | Covid 19 vaccine uptake |  |  |  | P-value |
| --- | --- | --- | --- | --- | --- |
|  | No hesitancy<br>341 (27.4%) | Acceptors with doubts<br>180 (14.5%) | Delayers<br>462 (37.1%) | Refusers<br>261 (21%) |  |
| Collective responsibilities |  |  |  |  |  |
| I wear face mask | 287 (84.2%) | 118 (65.7%) | 270 (58.4%) | 144 (55.2%) | <0.01* |
| I keep physical distance | 135 (39.6%) | 45 (24.9%) | 102 (22.1%) | 9 (3.5%) | 0.63 |
| I wash hands in public place | 162 (47.5%) | 73 (40.6%) | 144 (31.2%) | 45 (17.2%) | 0.04* |
| I wash hands at home | 117 (34.3%) | 59 (32.8%) | 106 (22.9%) | 36 (13.8%) | 0.06 |
| Take collective responsibility |  |  |  |  |  |
| Agree | 284 (83.3%) | 73 (40.6%) | 144 (31.2%) | 46 (17.6%) | 0.02* |
| Disagree | 57 (16.7%) | 107 (59.4%) | 318 (68.8%) | 215 (82.4%) |  |
| Convenience |  |  |  |  |  |
| I don't have time | - | 17 (9.4%) | 53 (11.5%) | 11 (4.2%) | 1.33 |
| Distance is far | - | 51 (28.3%) | 191 (41.3%) | 87 (33.3%) | 0.04* |
| Poor quality of healthcare | - | 3 (1.7%) | 11 (2.4%) | - | 0.87 |
| **Complacency** |  |  |  |  |  |
| Know much information about Covid 19 | 122 (35.7%) | 137 (76%) | 275 (59.5%) | 93 (35.6%) | 0.02* |
| Read negative information | 138 (40.4%) | 161 (89.4%) | 440 (95.2%) | 242 (92.7%) | <0.01* |
| Opinion of family and friends is important | 129 (37.9%) | 115 (63.9%) | 262 (56.7%) | 99 (37.9%) | 0.02* |
| Opinion of healthcare workers is not important | 144 (42.2%) | 118 (65.6%) | 274 (59.3%) | 110 (42.1%) | 0.01 * |
| I don't think I will be infected | 36 (10.6%) | 41 (22.8%) | 97 (21%) | 102 (39.1%) | 0.03* |
| I am against vaccines in general | 7 (2.1%) | 34 (18.9%) | 462 (100%) | 261 (100%) | 0.56 |
| I fear injections | 39 (11.4%) | 48 (26.7%) | 131 (28.4%) | 44 (16.9%) | 0.48 |
| Vaccine is not important | 114 (33.4%) | 115 (63.6%) | 195 (42.3%) | 87 (33.3%) | 0.02* |
| Complacent to receive vaccine | 124 (36.4%) | 137 (76.1%) | 395 (85.5%) | 234 (89.7%) | 0.04* |
| Agree | 217 (63.6%) | 43 (23.9%) | 67 (14.5%) | 27 (10.4%) |  |
| Disagree |  |  |  |  |  |

\* Indicates statistical significance ( $p < 0.05$ )

Collective responsibility was highly associated with no hesitancy (83.3%), with wearing a face mask and washing hands in public places being the only significant parameters. Acceptance with doubts and intentions to delay or refuse the vaccine were negatively associated with the distance from one’s home to the Covid-19 vaccination center.

A significant proportion of those who expressed some degree of complacency towards the Covid-19 vaccine were more likely to accept the vaccine with doubts (76.1%), delay (85.5%), or refuse (89.7%) it. This was particularly true among those who knew a lot of information about the vaccine, whose opinion of family and friends was important, and who thought they would not get infected. Those who read negative information or had bad experiences or knowledge of someone with a bad experience also expressed some degree of doubt and intention to delay or refuse the vaccine.

### Determinants of Covid 19 vaccine hesitancy

Multinomial logistic regression was used to analyze the determinants of Covid-19 vaccine uptake, with no hesitancy as the reference category (Table 4). The results showed that respondents aged below 70 years were more likely to accept the vaccine with doubts (AOR = 16.4; 95% CI = 15.92-20.76), while those aged 80 years were less likely to accept the vaccine with doubts (AOR = 0.33; 95% CI = 0.06-0.49) compared to their older counterparts. Level of education was significantly associated with Covid-19 vaccine uptake, with respondents with secondary education having higher odds of accepting the vaccine with doubts (AOR = 2.99; 95% CI = 2.07-4.18) or refusing it altogether (AOR = 4.10; 95% CI = 3.72-6.45) compared to the reference category. Post-secondary education was significantly associated with higher odds of accepting the vaccine with doubts (AOR = 2.11; 95% CI = 1.76-2.80), delaying (AOR = 3.13; 95% CI = 1.91-4.15), or refusing (AOR = 3.02; 95% CI = 2.47-4.38) it. Respondents with chronic diseases had higher odds (AOR = 2.12; 95% CI = 1.53-3.37) of accepting the Covid-19 vaccine compared to the no hesitancy reference group. Trust in decision makers was significantly associated with a higher likelihood of refusing the Covid-19 vaccine (AOR = 2.59; 95% CI = 2.31-3.04). Collective responsibility was negatively associated with the likelihood of accepting the vaccine with doubts (AOR = 4.12; 95% CI = 3.76-4.91), delaying (AOR = 0.05; 95% CI = 0.04-0.06), or refusing (AOR = 0.30; 95% CI = 0.25-0.40) it. Elderly respondents who had to travel long distances were more likely to delay their first Covid-19 vaccination dose (AOR = 2.64; 95% CI = 1.62-4.71). Complacency was also significantly associated with the intention to delay (AOR = 1.83; 95% CI = 1.30-2.31) or refuse (AOR = 3.40; 95% CI = 2.98-4.30) the Covid-19 vaccine. Marital status, primary occupation, and trust in vaccine safety and delivery system were not significantly associated with the uptake of the Covid-19 vaccine.

**Table 4:** Determinants of Covid 19 vaccine uptake by multinomial logistic regression (no hesitancy as reference category)

| **Variables** | **Acceptors with doubts** | **Delayers** | **Refusers** |
| --- | --- | --- | --- |
| **Age category** |  |  |  |
| <70 years | 16.4 (15.92, 20.76) * | 1.97 (1.22, 2.56) | 3.06 (2.23, 4.19) |
| 70 to 80 years | 3.52 (2.86, 6.04) | 0.72 (0.09, 1.62) | 3.50 (2.41, 5.56) |
| >80 years | 0.33 (0.06-0.49) ** | 1.63 (0.44, 2.51) | 1.72 (0.89, 4.38) |
| **Marital status** |  |  |  |
| Single | 3.7 (2.88, 6.31) | 3.89 (2.23, 5.19) | 1.33 (0.31, 2.07) |
| Married | 0.15 (0.07, 0.34) | 3.98 (1.37, 6. 55) | 2.0 (0.43, 3.61) |
| Divorced | 1.89 (0.61, 5.85) | 1.82 (0.36, 7.13) | 3.71 (0.41, 5.56) |
| **Level of education** |  |  |  |
| Primary | 0.32 (0.76, 1.32) | 0.53 (0.15, 1.92) | 0.33 (0.09, 1.24) |
| Secondary | 2.99 (2.07, 4.18) * | 0.36 (0.08, 1.63) | 4.10 (3.72, 6.45) * |
| Post-secondary | 2.11 (2.01, 2.80) * | 3.13 (1.91, 4.15) * | 3.02 (2.47, 4.38) * |
| None | 1.04 (0.23, 5.33) | 0.72 (0.41, 1.01) | 0.48 (0.27, 1.53) |
| **Primary occupation** |  |  |  |
| Farming | 0.88 (0.18, 4.42) | 0.08 (0.01, 2.42) | 0.82 (0.15, 2.41) |
| Business | 0.91 (0.14, 6.04) | 0.53 (0.11, 2.55) | 0.34 (0.03, 3.69) |
| Informal | 1.34 (0.89, 5.76) | 0.57 (0.09, 3.73) | 1.59 (0.36, 5.10) |
| **Chronic disease** | 2.12 (1.53-3.37) ** | 0.57 (0.19, 1.71) | 2.48 (0.61, 4.06) |
| Trust vaccine is safe for use | 1.06 (0.29, 3.86) | 3.77 (3.35, 4.12) | 0.35 (0.31, 0.38) |
| Trust in decision makers | 1.89 (1.62, 2.11) | 3.47 (2.83, 3.65) | 2.59 (2.31, 3.04) ** |
| Trust in delivery system | 3.57 (1.63, 7.32) | 2.00 (1.71, 2.38) | 2.83 (2.47, 2.97) |
| Take collective responsibility | 4.12 (3.76, 4.91) * | 0.05 (0.04, 0.06) * | 0.30 (0.25, 0.40) * |
| Distance is far | 2.64 (1.62, 4.71) * | 1.38 (0.72, 3.69) | 4.05 (2.66, 5.33) |
| Complacent to receive vaccine | 0.93 (0.56, 2.23) | 1.83 (1.30, 2.31) * | 3.40 (2.98, 4.30) ** |

\* Indicates statistical significance ( $p < 0.05$ )
\*\* Indicates statistical significance ( $p < 0.01$ )

## Discussion

### Socio-demographic characteristics

In this study, we found that age, education, and source of income were key determinants of vaccine acceptance and hesitancy among the elderly population in Kenya. Previous research has shown that younger age, lower education, and lower income are often associated with vaccine hesitancy (12, 13). However, in our study, we found that being below the age of 70 was a predictor of vaccine acceptance with doubts, but not resistance or delay. In contrast, being above the age of 80 was associated with a decrease in the odds of hesitancy. This may be due to the fact that the elderly population, who are at the greatest risk of adverse Covid-19 disease outcomes (14, 15), were intentionally targeted for vaccination.

We also found that education was significantly associated with vaccine uptake, with those with secondary or post-secondary education more likely to be classified as acceptors with doubts. This may be because the most educated in a population group are often the major users of health services and are more able to understand health promotional messages (16, 17). However, in our study, the most educated were more likely to hesitate taking the vaccine. This could be due to the type of information they assessed, as well as their high trust in the opinions of family and friends and political leaders for decision-making. Previous studies showed association between hesitancy toward vaccines and misconceptions about vaccinations among certain populations (18, 19, 20).

To improve vaccine uptake in our study population, it may be necessary to focus on health promotion efforts that target specific context-specific messages to clarify the truth and inform the public about the safety and importance of the vaccine. Given the high levels of trust in health workers among our respondents, this may be most effectively done through intentional health promotion efforts by health workers. The goal should be to provide the right message at the right time to ensure that the public is accurately informed about the Covid-19 vaccine.

### Confidence in Covid 19 vaccine

Several studies conducted in western populations have reported an association between a short development time for a vaccine and vaccine uptake (21, 22). However, the present study did not find such a relationship. Instead, fear that the vaccine contained harmful substances was significantly associated with distrust in the vaccine’s safety (23). This finding confirms an earlier report by (24), which showed an increased odds of vaccine refusal among the African population due to misconceptions about the vaccine’s contents. While there are misconceptions about the content of the vaccine (25), government officials have consistently educated the public about the development process for the Covid-19 vaccine (26). Therefore, distrust in the vaccine’s safety is a cause for concern given the extensive sensitization efforts by both the county and national governments and the high number of respondents who had a lot of information about the Covid-19 vaccine.

The source and type of information provided are important factors in facilitating behavior change and acceptance of intervention processes and outcomes (27). Similarly, studies by (28) have reported increased utilization of vaccination services among the most informed population groups, leading to low vaccine hesitancy. They reported high acceptance of measles and tetanus vaccines among population groups who received information from public health officers, nurses, and community health workers, with local vernacular radio stations as the medium of delivery. Our findings are consistent with these reports, but it is important to consider the roles played by other factors in the 5Cs model in enabling vaccine acceptance (26).

Severe Covid-19 disease and worse outcomes have been associated with comorbidities, particularly among cases with Delta and Kappa coronavirus strains (29). A high case fatality rate (40%) has been reported among Covid-19 patients with diabetes and cardiovascular diseases (26). In the present study, a significant proportion of respondents with comorbidities were diabetic and were more likely to hesitate taking the vaccine. If previous reports are to be believed, this hesitation may be due to misconceptions and negative information about severe reactions to the vaccine and the harmful substances they contain, leading to fear of worse health outcomes (28). While there are limited studies in this area, it is not clear that increased vaccine uptake would lead to higher fatality rates or adverse health outcomes. However, if the rare cases of health outcomes after receiving the first or second jab of vaccines are considered, there is a potential for a reaction to the vaccine, regardless of the type of vaccine or manufacturer (21). Further studies in this emerging area are recommended, as it was outside the scope of our study.

### Other 5Cs of vaccine hesitancy

Complacency to vaccine can be prevented through targeted health promotion, such as by providing education and awareness campaigns (30, 31). This leads to positive perception about the vaccine and raises trust in the importance of the vaccine to the population (32, 33). Past studies have reported bad experience with the previous vaccines, general lack of safety of vaccines, and lack of vaccine compatibility with personal and religious beliefs as the key pointers of complacency (31, 34). In the current study, these didn’t concern the respondents, instead they were generally against Covid 19 vaccine and could not relate with its importance (34). Other reasons in our study which contributed to complacency and are previously reported included perception that one is not likely to be infected with the disease, reading negative information and general feeling of having enough information to keep one safe (33, 34). Additionally, past studies showed negative association between complacency and collective responsibilities (30). Threefold likelihood of complacency was observed among the respondents that had recommendable scores for collective responsibilities (35). The findings for the present work were in tandem with these earlier reports (36, 37, 38, 39, 40). However, in order to make a conclusive decision, more work need to be done in this area.

In developed countries not having time was the main reported reason for lack of convenience (41). Similar studies in developing countries have reported two major reasons for lack of convenience in Covid 19 vaccine to be long distance from the vaccination centers and homesteads and lack of knowledge about the vaccinations (42, 43). Our study agreed with these findings since long distance was associated with nearly threefold likelihood of hesitancy. Indeed, earlier reports showed low child immunization rates among caregivers living several miles away from health facilities and among the elderly (44, 45, 46). In addition, lack of knowledge about the vaccine was associated with increased likelihood of hesitancy in both our study and earlier studies (47, 48, 49). Therefore, it is evident from our findings and earlier studies that lack of convenience and lack of knowledge about the vaccine are major reasons for hesitancy. To reduce the hesitancy, it is important for the government to ensure easy access to the vaccine and to provide comprehensive information about the vaccine to the public. Community-based awareness campaigns should be encouraged in order to ensure that the people are better informed about the benefits of the vaccine and its safety. In addition, governments should also focus on making the vaccine more convenient by setting up more vaccination centers and providing transportation to those who are far from the centers.

## Conclusion

In conclusion, our study found that age, education, source of income, and other socio-demographic characteristics were significant factors in vaccine acceptance and hesitancy among the elderly population in Kenya. Fear that the vaccine contained harmful substances was significantly associated with distrust in the vaccine’s safety, while lack of knowledge about the vaccine and lack of convenience were associated with hesitancy. These findings indicate the need for health promotion efforts that target specific context-specific messages to clarify the truth and inform the public about the safety and importance of the vaccine. In addition, governments should focus on making the vaccine more accessible and convenient through more vaccination centers and providing transportation to those who are far from the centers. Furthermore, comprehensive information about the vaccine should be provided to the public to ensure that they are better informed about the benefits of the vaccine and its safety. In order to achieve the desired outcomes, it is essential that all stakeholders actively collaborate to ensure that the best strategies are implemented in order to encourage the elderly population to accept and receive the Covid-19 vaccine.

## Data Availability

All data are provided as part of the manuscript.

## Acknowledgment

The authors would like to acknowledge the contributions of all those who were involved in the completion of this work. We are grateful to the participants who generously gave their time and insights, as well as the research team who conducted the study. We would also like to thank our colleagues and mentors who provided guidance and support throughout the research process. Finally, we would like to express our appreciation to the members of the research team who pulled together their resources to fund the project, kudos.

